# CAUSAL-RSV: A study protocol for a causal mediation analysis of RSV vaccine effects in infants using real-world data

**DOI:** 10.64898/2026.07.16.26356876

**Authors:** Annette K. Regan, Matthew M. Coates, Sheena G. Sullivan, Flor M. Muñoz, Stacey L. Rowe, Chantal Avila, Onyebuchi A. Arah

## Abstract

Respiratory syncytial virus (RSV) contributes to substantial morbidity and mortality in young children each year. In 2023, two new prevention products were licensed and recommended in the United States (US), including a prefusion F protein subunit vaccine (RSVpreF) administered during pregnancy and a long-acting monoclonal antibody (mAb) administered in infants.

Although post-licensure real-world studies support the effectiveness of RSVpreF vaccine during pregnancy, existing studies have been conducted in settings where only RSVpreF vaccine is available. The real-world effectiveness of RSVpreF vaccine in settings where both RSVpreF vaccine and mAbs are available is not yet well understood. The goal of this study is to estimate the real-world effectiveness of the RSVpreF vaccine against severe infant RSV by applying causal mediation analysis with receipt of mAbs as a mediating variable. Using a national cohort of mother-infant dyads with the Optum Labs Data Warehouse (OLDW), we will model vaccine and mAb effects in a longitudinal cohort spanning the 2023-24, 2024-25, and 2025-26 RSV seasons. Results will be used to better understand the total effect of RSVpreF vaccination when it is used as one component within a hybrid infant RSV prevention program.

## 2. RATIONALE AND BACKGROUND

Respiratory syncytial virus (RSV) is the leading cause of hospitalization in young children, contributing to 33 million episodes of RSV-associated acute lower respiratory tract disease (LRTD), 3.6 million RSV-associated LRTD hospitalizations, and 101,400 RSV-attributable deaths in children aged <5 years annually.^1^ RSV is a common illness in children aged <5 years, with 15% of children experiencing a medically-attended RSV-associated LRTD at any time in the first five years of life.^2,3^ RSV contributes substantially to pediatric morbidity and mortality globally, with one in every 50 deaths in children aged <5 y attributed to RSV infection.^1^ The burden is especially prominent in early infancy, with the highest incidence of severe illness occurring in infants aged <3m. More than 50% of infections occur in the first year of life,^2^ and one in every 28 deaths among infants aged <6m can be attributed to RSV infection.^2,3^ Although several known risk factors such as prematurity, chronic lung disease, congenital heart disease, and other pre-existing medical conditions can place children at higher risk of severe illness, 50-95% of RSV hospitalizations occur in otherwise healthy children with no known comorbidities.^1^

Treatment and prevention options for RSV have been historically limited. Although the monoclonal antibody (mAb) palivizumab has been available in the US for RSV prophylaxis since 1998, the American Academy of Pediatrics (AAP) has previously recommended palivizumab only for children with underlying medical conditions.^4,5^ Palivizumab is an expensive treatment with a monthly dosing schedule,^4^ and is therefore not cost-effective for healthy children,^6^ who make up the bulk of hospitalized RSV cases.^3^ In July 2023, the US Food and Drug Administration (FDA) licensed a novel long-acting mAb product, nirsevimab (Beyfortus^®^), to provide passive immunization to prevent severe RSV LRTD in all infants during their first RSV season and children with underlying medical conditions in their second RSV season.^7^ In June 2025, a second long-acting mAb was licensed by the FDA (clesrovimab, Enflonsia^®^) with a similar indication, with exception to second season administration for children with underlying medical conditions.^8^ Both products are recommended for seasonal administration by the US Advisory Committee on Immunization Practices (ACIP)^9,10^ and the American Academy of Pediatrics (AAP)^11^and the American College of Obstetricians and Gynecologists (ACOG)^12^ for infants 0-8 months old during the RSV season.

While long-acting mAb products are offered near or after birth, maternal vaccination can offer passive immunity from birth via transplacental transferred maternal antibodies. In 2023, the US FDA licensed a bivalent RSV prefusion F protein-based (RSVpreF) vaccine for administration during pregnancy to prevent RSV illness in both healthy and high-risk children.^13^ In September 2023, the US ACIP recommended one dose of RSVpreF vaccine for pregnancies at 32-36 weeks’ gestation between September and January to prevent severe RSV-associated LRTD among infants.^14^

Clinical studies have demonstrated that RSVpreF vaccination during pregnancy produces maternal antibodies, which cross the placenta and offer infants protection from birth.^15,16^ Results from a phase 2b clinical trial demonstrated that when administered between 24 and 36 weeks of pregnancy, the vaccine elicits neutralizing antibodies in maternal serum at delivery and in cord blood.^16^ Transplacental neutralizing antibody transfer ratios ranged from 1.4 to 2.1 and this ratio was consistent across different gestational ages at administration.^16^ Subsequent data from the MATISSE trial, a phase 3 double-blind clinical trial conducted in 18 countries, showed that RSVpreF vaccination between 24 and 36 weeks of pregnancy resulted in 81% fewer medically-attended severe LRTD, 57% fewer RSV-associated LRTD, and 68% fewer RSV-associated hospitalizations among infants aged <3 months.^17^ By age 6m, infants born to vaccinated mothers had 69% fewer medically-attended severe LRTD, 51% fewer RSV-associated LRTD, and 57% fewer RSV-associated hospitalizations.^17^

In addition to prelicensure clinical trials and modeling studies,^6,18,19^ post-licensure real-world studies support the effectiveness of RSVpreF vaccination during pregnancy in preventing RSV-associated LRTD among infants. A nested case-control study in Scotland showed that RSVpreF vaccination from 28 weeks of gestation was 82.2% effective against RSV-related LRTD, and these results were similar when infants were born preterm or term.^20^ Another test-negative design study in the UK (Scotland and England) showed RSVpreF vaccination from 28 weeks of gestation was 72% effective when the vaccine was given more than 14 days before delivery.^21^ In Argentina, two separate test-negative design studies showed that vaccination between 32 and 36 weeks of pregnancy was 77-79% effective against severe RSV-associated LRTD.^22,23^

While these studies support the real-world effectiveness of RSVpreF vaccine, these studies were conducted in settings with only RSVpreF availability. The effects of RSVpreF vaccine in settings where both RSVpreF vaccine and mAbs are available are not yet well understood. One French cohort study directly compared the effectiveness of mAb versus RSVpreF administration, showing that nirsevimab was modestly more effective than RSVpreF vaccination during pregnancy; nirsevimab conferred a 26% relative reduction in RSV-associated LRTD in comparison to RSVpreF vaccine and a 42% relative reduction in severe outcomes, including pediatric intensive care unit admission, ventilator support, or oxygen therapy.^24^

## 3. RESEARCH QUESTION AND OBJECTIVES

Our overall goal is to estimate the real-world effectiveness of the RSVpreF vaccine by applying an innovative causal inference framework to a large, administrative healthcare database. We will use national claims data to evaluate the performance of RSVpreF vaccine across the 2023-24, 2024-25, and 2025-26 RSV seasons in the US, during periods of mAb availability. To achieve these goals, we will address the following objectives:

### Objective 1. Estimate the total direct and indirect real-world effects of maternal RSVpreF vaccination against infant RSV-attributable illness and hospitalization

We will conduct a causal mediation analysis of time-to-event outcome models to estimate the total effect of RSVpreF vaccination on infant RSV illness, including the controlled direct and pure indirect effects. We will estimate the direct effect of RSVpreF vaccine on infant illness and the indirect effect of RSVpreF vaccine on infant illness via the reduced probability of nirsevimab administration. We hypothesize that both effects are important for explaining the infant health effects of maternal RSVpreF vaccine in settings with mAb recommendations.

### Objective 2. Quantify the heterogeneity in the total real-world effectiveness of maternal RSVpreF vaccination by gestational age at vaccination and high-risk groups

In our secondary objective, we will extend our causal mediation analysis to estimate the direct and indirect effects of RSVpreF vaccination by gestational age at vaccination and by high-risk group, including pregnancies with health complications and infants with high-risk conditions for severe RSV illness. We hypothesize that the direct and indirect effects of RSVpreF vaccine will vary by gestational age at vaccination and the presence of high-risk conditions.

## 4. RESEARCH METHODS

### 4.1 Study Design

A national cohort study will be conducted using the Optum Labs Data Warehouse (OLDW). Using available de-identified claims information, we will construct a longitudinal cohort of mother-infant pairs with an infant date of birth between 2023 and 2025. Pregnancies will be identified using a previously validated claims-based algorithm.^25^ In brief, the algorithm assigns a pregnancy outcome and pregnancy start date (i.e., estimated last menstrual period [LMP]) based on available pregnancy-related medical encounters.^25^ Infants can be linked to mothers based on family identifiers associated with both the mother and infant’s healthcare plan.

Mother-infant pairs will be eligible for inclusion if they meet the following criteria:

1. The mother is aged 18-49 years old at LMP;
2. The infant is live born;
3. The mother has a pregnancy overlapping RSV vaccine eligibility (see Figure S1);
4. The infant is either born during the RSV season or is aged <8 months at the start of the RSV season (see Figure S1);
5. The infant is not exposed to palivizumab (see Table S1);
6. The mother is continuously enrolled for care for 365 days preceding LMP (to ensure capture of baseline covariates).

We will establish start and end dates for the RSV season for the 2023-24, 2024-25, and 2025-26 seasons using the weekly percent of tests positive for RSV reported to the National Respiratory and Enteric Virus Surveillance System (NREVSS).^26^ Based on prior work from the Centers for Disease Control and Prevention (CDC), we identified the start of the season as the second consecutive surveillance week with ≥3% of tests positive for RSV and the end of the season as the second consecutive surveillance week with <3% of tests positive for RSV.^27^

For eligible mother-infant pairs, we will extract all available inpatient and outpatient claims, laboratory testing records (and results), and immunization procedures to define study measures (Figure).

**Figure.**
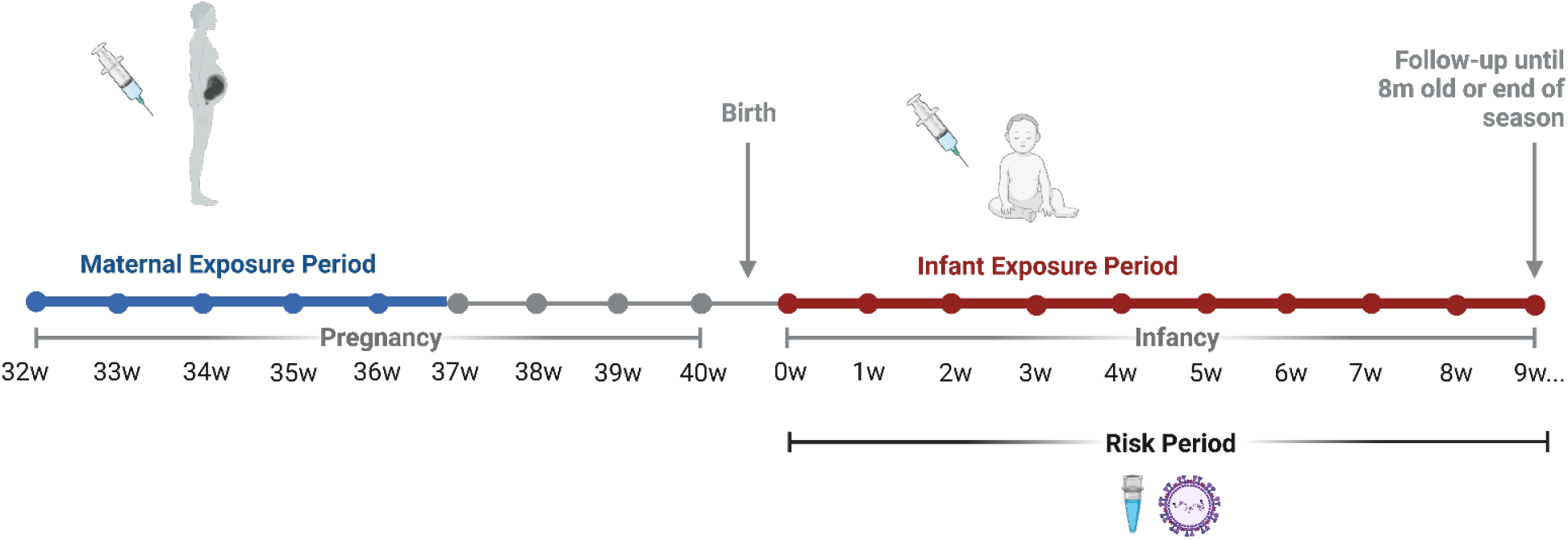
Exposure and mediator measurement and risk period for a study of maternal RSVpreF vaccination and infant RSV illness.

### 4.2 Data Sources

The OLDW is a large-scale, de-identified, longitudinal dataset that integrates medical, pharmacy, and laboratory claims with enrollment records, covering over 350 million de-identified individuals enrolled in either commercial insurance or Medicare Advantage plans across the US.^28,29^ It is widely used in health services and research.^29^ A limited, de-identified OLDW dataset is made available to study team members from UCLA for the purpose of research for a two-year period using a Virtual Data Interface. All data are statistically de-identified to protect privacy.

### 4.3 Exposure and mediator definitions

We will examine all maternal inpatient and outpatient records for evidence of RSVpreF vaccination during pregnancy (exposure variable, Table S2) and the date and gestational age at vaccination during the maternal exposure period using current procedural terminology (CPT) codes and national drug codes (NDC, see Table S1 for codes).

Similarly, we will examine all infant inpatient and outpatient claims records for evidence of nirsevimab or clesrovimab administration (mediating variable) among infants <8 months old.

### 4.4 Outcome definitions

Using International Classification of Diseases, Tenth Revision, Clinical Modification (ICD-10-CM) codes in outpatient and inpatient medical claims records and Logical Observation Identifiers Names and Codes (LOINC) codes in linked laboratory testing records (Table S3) during the infant risk period (i.e., birth to <8 months of age), we will identify all medically-attended RSV-associated LRTD hospitalizations (Table S4), defined as an inpatient claim with an LRTD diagnosis (see Table S5 for codes). This outcome aligns with those used by the pre-licensure clinical trials.^5 21^ To evaluate the sensitivity of our outcome in detecting severe RSV, we will additionally evaluate (1) the proportion of medically-attended RSV-associated LRTD hospitalizations requiring mechanical ventilation and/or critical care (i.e., severe medically-attended RSV-associated LRTD; (2) the proportion of all-cause LRTD with a laboratory RSV detection.

### 4.5 Covariates

Using enrollment data, pharmaceutical claims, and outpatient and inpatient medical claims records, we will identify maternal age, household income, educational attainment, comorbidities, pregnancy complications, and receipt of other vaccines recommended during pregnancy (i.e., Tdap, influenza, COVID-19) (Table S6, see Table S7 for codes). Infant covariates will include infant sex, prematurity, low birthweight, small-for-gestational age birth, mode of delivery, admission to neonatal intensive care unit, and underlying health conditions (i.e., Trisomy 21, chronic lung disease of prematurity, congenital heart disease, neuromuscular disease impairing respiratory function, and immunocompromising conditions) (see Table S8 for codes).

### 4.6 Statistical analysis

#### 4.6.1 Analysis of RSVpreF vaccine and monoclonal antibody administration

Within mother-infant dyads, we will evaluate patterns of RSVpreF vaccine administration during pregnancy among mothers in the study cohort and receipt of mAbs during the first 8 months of life among infants in the study cohort. We will calculate the proportion of dyads who received (1) RSVpreF vaccine only, (2) mAb only, (3) both a RSVpreF vaccine and mAb, or (4) neither RSVpreF vaccine nor mAb. Dyads in which the RSVpreF vaccine is administered outside of the recommended 32-36 week window will be excluded. We anticipate the number of these dyads to be small. We will use descriptive statistics to examine administration patterns over time and by maternal and infant characteristics. We will quantify differences in the odds of administration patterns using multinomial logistic regression, with no RSVpreF vaccine or mAb as the reference.

#### 4.6.2 Mediation analysis

For causal mediation analyses, we will treat maternal RSVpreF vaccination as our exposure of interest (*X*), infant receipt of an mAb product as the mediator (*M*), and medically-attended RSV-associated LRTD as the outcome (*Y*). Models will account for maternal (*C*_*M*_) and infant covariates (*C*_*I*_). We will estimate the mediator model as:

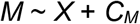

to examine how the exposure affects the mediator. We will estimate the outcome model as:

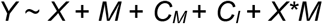

to examine how the mediator and exposure affect the outcome, allowing for exposure-mediator interaction. We will conduct a four-way decomposition^30^ of the total effect into the direct and indirect effects. We will estimate causal effects by modeling a single exposure and multiple sequential mediators, which can be fit as a time-varying mediator (e.g., mediator = c(“M_t1”,”M_t2”,”M_t3”)). Because maternal RSVpreF vaccination and infant administration of monoclonal antibodies both confer passive immunity against RSV through related biological mechanisms, we will consider the possibility of an exposure–mediator interaction, allowing for interaction between maternal vaccination and mAb receipt in the outcome model.

#### 4.6.3 Heterogeneity analysis

If sufficient data are available, we will evaluate whether heterogeneity of effects by estimating separate mediation effects in stratified models by gestational week of vaccination (32 weeks, 33 weeks, 34 weeks, 35 weeks, or 36 weeks), by prematurity (preterm or term), and presence of an underlying health condition (yes or no). To estimate whether effects statistically vary, we will fit an interaction term by gestational week, prematurity, and presence of an underlying health condition.

### 4.7 Data quality review

We will examine the proportion and distribution of missing variables by maternal and infant covariates. To evaluate the validity of data variables, we will estimate the prevalence of exposures, covariates, and outcome variables and compare these to published US estimates. Depending on the results of these checks, we may revise selected ICD-10-CM and other codes for the identification of variables. Quantitative bias analyses will be used to evaluate the potential influence of exposure and outcome misclassification based on these data quality checks. E values may be used to evaluate the influence of unmeasured confounding variables on study findings.

## Data Availability

There are no data associated with this submission.

## 5. Institutional Review Board Approval

This protocol was reviewed by the Institutional Review Board of the University of California Los Angeles and determined to not involve research with human subjects as defined by the Department of Health & Human Services (DHHS) and Food and Drug Administration (FDA) regulations. IRB review and approval was therefore not required (IRB-25-2622).

## 6. Plans for Disseminating and Communicating Results

Manuscripts, abstracts, press releases and other publications detailing the study’s procedures or findings will be prepared collaboratively by the study team. The study team will follow the International Committee of Medical Journal Editors (ICMJE) criteria to determine authors, and all authors who meet these criteria will be offered authorship. Publications will include a list of investigators, with authors being determined in line with the ICMJE guidelines.

## LIST OF ABBREVIATIONS

**Abbreviation Definition**

AAP: American Academy of Pediatrics
ACIP: Advisory Committee on Immunization Practices
ACOG: American College of Obstetricians and Gynecologists
ARI: Acute Respiratory Illness
CDC: Centers for Disease Control and Prevention
CMAverse: Causal Mediation Analysis in R
COVID-19: Coronavirus Disease of 2019
CPT: Current Procedural Terminology
DHHS: Department of Health & Human Services
EHR: Electronic Health Record
FDA: Food and Drug Administration
GA: Gestational Age
HELLP: Hemolysis, Elevated Liver Enzymes, Low Platelet Count
ICD-10-CM: International Classification of Diseases, Tenth Revision, Clinical Modification
ICMJE: International Committee of Medical Journal Editors
IRB: Institutional Review Board
LMP: Last Menstrual Period
LOINC: Logical Observation Identifiers, Names, and Codes
LRTD: Lower Respiratory Tract Disease
mAb: Monoclonal Antibody
NDC: National Drug Codes
NICU: Neonatal Intensive Care Unit
NREVSS: National Respiratory and Enteric Virus Surveillance System
OLDW: Optum Labs Data Warehouse
PCR: Polymerase Chain Reaction
RSV: Respiratory Syncytial Virus
RSVpreF: RSV Prefusion F Protein-based Vaccine
RT-PCR: Real-time Polymerase Chain Reaction
Tdap: Tetanus, Diphtheria and Acellular Pertussis Vaccine
UCLA: University of California Los Angeles
UK: United Kingdom
US: United States

## 7. Data Availability Statement

The study is conducted under a data use agreement between the study team and Optum Labs. The authors do not have permission to share study data with other researchers. Permission to access data must be requested to Optum Labs.

## 8. Competing Interests

Annette K. Regan reports membership on a data safety monitoring board (DSMB) for matters unrelated to the presented research, consulting fees for unrelated research from the Pan American Health Organization, and unrelated research funding paid to her institution from Merck Sharp & Dohme, Pfizer, and Bavarian Nordic. Sheena G. Sullivan reports consulting for Astra-Zeneca, GSK, Sanofi, Pfizer, Moderna, CSL Seqirus, Novavax, and Evo Health on work unrelated to the presented research. Flor M. Munoz has been an investigator in pediatric studies of coronavirus disease 2019 vaccines for Pfizer and Moderna, and for a pediatric remdesivir study conducted by Gilead Sciences; is an investigator on projects supported by the National Institutes of Health (NIH) and the Centers for Disease Control and Prevention (CDC); has served as a member of the DSMB for clinical trials conducted by Pfizer, Dynavax, Moderna, Meissa Vaccines, Virometix, and the NIH; and has served as a consultant to Merck, Moderna, Sanofi, Astra Zeneca, and GSK. Stacey L. Rowe reports consulting for CSL Seqirus on work unrelated to the presented research. The other authors do not report any potential conflicts of interest.

## 9. Funding

Research reported in this publication was supported by the National Institute of Allergy and Infectious Diseases of the National Institutes of Health under Award Number R21AI191038. The content is solely the responsibility of the authors and does not necessarily represent the official views of the National Institutes of Health.

## References

1. Li Y, Wang X, Blau DM, et al. Global, regional, and national disease burden estimates of acute lower respiratory infections due to respiratory syncytial virus in children younger than 5 years in 2019: a systematic analysis. Lancet Lond Engl. 2022;399(10340):2047–2064. doi:10.1016/S0140-6736(22)00478-0

2. Hall Caroline Breese, Weinberg Geoffrey A., Iwane Marika K., et al. The Burden of Respiratory Syncytial Virus Infection in Young Children. N Engl J Med. 360(6):588–598. doi:10.1056/NEJMoa0804877

3. Suss RJ, Simões EAF. Respiratory Syncytial Virus Hospital-Based Burden of Disease in Children Younger Than 5 Years, 2015-2022. JAMA Netw Open. 2024;7(4):e247125–e247125. doi:10.1001/jamanetworkopen.2024.7125

4. Viguria N, Navascués A, Juanbeltz R, Echeverría A, Ezpeleta C, Castilla J. Effectiveness of palivizumab in preventing respiratory syncytial virus infection in high-risk children. Hum Vaccines Immunother. 2021;17(6):1867–1872. doi:10.1080/21645515.2020.1843336

5. Jones JM, Fleming-Dutra KE, Prill MM, et al. Use of Nirsevimab for the Prevention of Respiratory Syncytial Virus Disease Among Infants and Young Children: Recommendations of the Advisory Committee on Immunization Practices - United States, 2023. MMWR Morb Mortal Wkly Rep. 2023;72(34):920–925. doi:10.15585/mmwr.mm7234a4

6. Yu T, Padula WV, Yieh L, Gong CL. Cost-effectiveness of nirsevimab and palivizumab for respiratory syncytial virus prophylaxis in preterm infants 29–34 6/7 weeks’ gestation in the United States. Pediatr Neonatol. 2024;65(2):152–158. doi:10.1016/j.pedneo.2023.04.015

7. FDA. FDA Approves New Drug to Prevent RSV in Babies and Toddlers. July 17, 2023. Accessed March 10, 2026. https://www.fda.gov/news-events/press-announcements/fda-approves-new-drug-prevent-rsv-babies-and-toddlers

8. AAP. FDA approves new monoclonal antibody to protect infants from RSV. June 10, 2025. Accessed March 10, 2026. https://publications.aap.org/aapnews/news/32373/FDA-approves-new-monoclonal-antibody-to-protect?autologincheck=redirected

9. Jones JM, Fleming-Dutra KE, Prill MM, et al. Use of Nirsevimab for the Prevention of Respiratory Syncytial Virus Disease Among Infants and Young Children: Recommendations of the Advisory Committee on Immunization Practices - United States, 2023. MMWR Morb Mortal Wkly Rep. 2023;72(34):920–925. doi:10.15585/mmwr.mm7234a4

10. Moulia DL, Link-Gelles R, Chu HY, et al. Use of Clesrovimab for Prevention of Severe Respiratory Syncytial Virus-Associated Lower Respiratory Tract Infections in Infants: Recommendations of the Advisory Committee on Immunization Practices - United States, 2025. MMWR Morb Mortal Wkly Rep. 2025;74(32):508–514. doi:10.15585/mmwr.mm7432a3

11. AAP. Recommended Child and Adolescent Immunization Schedule for Ages 18 Years or Younger. Accessed June 25, 2026. https://downloads.aap.org/AAP/PDF/AAP-Immunization-Schedule.pdf

12. ACOG. 2026 Maternal Immunization Schedule. Accessed June 25, 2026. https://www.acog.org/clinical-information/maternal-immunization-schedule.

13. FDA. FDA Approves First Vaccine for Pregnant Individuals to Prevent RSV in Infants. Accessed February 23, 2026. https://www.fda.gov/news-events/press-announcements/fda-approves-first-vaccine-pregnant-individuals-prevent-rsv-infants

14. Fleming-Dutra KE, Jones JM, Roper LE, et al. Use of the Pfizer Respiratory Syncytial Virus Vaccine During Pregnancy for the Prevention of Respiratory Syncytial Virus-Associated Lower Respiratory Tract Disease in Infants: Recommendations of the Advisory Committee on Immunization Practices - United States, 2023. MMWR Morb Mortal Wkly Rep. 2023;72(41):1115–1122. doi:10.15585/mmwr.mm7241e1

15. Jasset OJ, Lopez Zapana PA, Bahadir Z, et al. Enhanced placental antibody transfer efficiency with longer interval between maternal respiratory syncytial virus vaccination and birth. Am J Obstet Gynecol. Published online November 7, 2024:S0002-9378(24)01125-6. doi:10.1016/j.ajog.2024.10.053

16. Bebia Z, Reyes O, Jeanfreau R, et al. Safety and Immunogenicity of an Investigational Respiratory Syncytial Virus Vaccine (RSVPreF3) in Mothers and Their Infants: A Phase 2 Randomized Trial. J Infect Dis. 2023;228(3):299–310. doi:10.1093/infdis/jiad024

17. Kampmann Beate, Madhi Shabir A., Munjal Iona, et al. Bivalent Prefusion F Vaccine in Pregnancy to Prevent RSV Illness in Infants. N Engl J Med. 2023;388(16):1451–1464. doi:10.1056/NEJMoa2216480

18. Averin A, Quinn E, Atwood M, Weycker D, Shea KM, Law AW. Cost-effectiveness of bivalent respiratory syncytial virus Prefusion F (RSVpreF) maternal vaccine among infants in the United States. Vaccine. 2025;58:127191. doi:10.1016/j.vaccine.2025.127191

19. Hutton DW, Prosser LA, Rose AM, et al. Cost-Effectiveness of Maternal Vaccination to Prevent Respiratory Syncytial Virus Illness. Pediatrics. 2024;154(6):e2024066481. doi:10.1542/peds.2024-066481

20. McLachlan I, Robertson C, Morrison KE, et al. Effectiveness of the maternal RSVpreF vaccine against severe disease in infants in Scotland, UK: a national, population-based case-control study and cohort analysis. Lancet Infect Dis. Published online November 28, 2025:S1473-3099(25)00624-3. doi:10.1016/S1473-3099(25)00624-3

21. Williams TC, Marlow R, Cunningham S, et al. Bivalent prefusion F vaccination in pregnancy and respiratory syncytial virus hospitalisation in infants in the UK: results of a multicentre, test-negative, case-control study. Lancet Child Adolesc Health. 2025;9(9):655–662. doi:10.1016/S2352-4642(25)00155-5

22. Gentile A, Juárez MDV, Lucion MF, et al. Maternal Immunization With RSVpreF Vaccine: Effectiveness in Preventing Respiratory Syncytial Virus-associated Hospitalizations in Infants Under 6 Months in Argentina: Multicenter Case-control Study. Pediatr Infect Dis J. 2025;44(10):988–994. doi:10.1097/INF.0000000000004878

23. Pérez Marc G, Vizzotti C, Fell DB, et al. Real-world effectiveness of RSVpreF vaccination during pregnancy against RSV-associated lower respiratory tract disease leading to hospitalisation in infants during the 2024 RSV season in Argentina (BERNI study): a multicentre, retrospective, test-negative, case-control study. Lancet Infect Dis. 2025;25(9):1044–1054. doi:10.1016/S1473-3099(25)00156-2

24. Jabagi MJ, Bertrand M, Gabet A, Kolla E, Olié V, Zureik M. Nirsevimab vs RSVpreF Vaccine for Respiratory Syncytial Virus-Related Hospitalization in Newborns. JAMA. 2026;335(9):787–798. doi:10.1001/jama.2025.24082

25. Moll K, Wong HL, Fingar K, et al. Validating Claims-Based Algorithms Determining Pregnancy Outcomes and Gestational Age Using a Linked Claims-Electronic Medical Record Database. Drug Saf. 2021;44(11):1151–1164. doi:10.1007/s40264-021-01113-8

26. CDC. The National Respiratory and Enteric Virus Surveillance System (NREVSS). Accessed February 24, 2026. https://www.cdc.gov/nrevss/php/dashboard/index.html

27. Hamid S, Winn A, Parikh R, et al. Seasonality of Respiratory Syncytial Virus - United States, 2017-2023. MMWR Morb Mortal Wkly Rep. 2023;72(14):355–361. doi:10.15585/mmwr.mm7214a1

28. Optum. Using real-world data to address the greatest challenges in healthcare. Accessed February 24, 2026. https://labs.optum.com/

29. Wallace PJ, Shah ND, Dennen T, Bleicher PA, Crown WH. Optum Labs: building a novel node in the learning health care system. Health Aff Proj Hope. 2014;33(7):1187–1194. doi:10.1377/hlthaff.2014.0038

30. Wang A, Arah OA. G-computation demonstration in causal mediation analysis. Eur J Epidemiol. 2015;30(10):1119–1127. doi:10.1007/s10654-015-0100-z

