## Supplemental Material for "CAUSAL-RSV: A study protocol for a causal mediation analysis of RSV vaccine effects in infants using real-world data"


Table S1. CPT and NDC codes for identification of RSVpreF vaccine, nirsevimab, and clesrovimab administration in administrative claims records……………………………………..….2

Table S2. Definition of exposure and mediator variables in CAUSAL-RSV analysis…………......3

Table S3. Logical Observation Identifiers Names and Codes (LOINC®) codes* for the identification of laboratory test results for respiratory syncytial virus……..…………………………..4

Table S4. Definition of primary and exploratory outcome variables in CAUSAL-RSV analysis.....5

Table S5. International Classification of Diseases (10th edition, clinical modification; ICD-10-CM) codes* to identify a diagnosis of lower respiratory tract disease…………………………………….6

Table S6. Definition of covariates in CAUSAL-RSV analysis……………………………………….7

Table S7. International Classification of Diseases (10th edition, clinical modification; ICD-10-CM), Current Procedural Terminology, and National Drug Codes to identify baseline maternal covariates………………………………………………………………..…………………………….9

Table S8. International Classification of Diseases (10th edition, clinical modification; ICD-10-CM) codes to measure infant health covariates………………………………………………………...12

Figure S1. Respiratory syncytial virus (RSV) activity used to establish maternal-infant eligibility criteria……………………………………………………………………………………………………14

**Table S1. CPT and NDC codes for identification of RSVpreF vaccine, nirsevimab, clesrovimab, and palivizumab administration in administrative claims records.**

| Variable | CPT* | NDC* | Description |
|---|---|---|---|
| RSVpreF vaccine | 90678 | 00069-0207-01<br>00069-0250-01<br>00069-0344-01<br>00069-2465-01<br>00069-2465-19 | Respiratory syncytial virus (RSV), vaccine, bivalent, protein subunit RSV prefusion F, diluent reconstituted, 0.5 mL, preservative free |
| Unspecified monoclonal antibody for RSV | 96380 | | Administration of respiratory syncytial virus, monoclonal antibody, seasonal dose by intramuscular injection, with counseling by physician or other qualified healthcare professional. |
| | 96381 | | Administration of respiratory syncytial virus, monoclonal antibody, seasonal dose by intramuscular injection. |
| Nirsevimab (Beyfortus®) | | 49281-0575-00,<br>49281-0575-15 | Respiratory syncytial virus (RSV) monoclonal antibody, IgG1κ, (nirsevimab-alip), 0.5 mL, neonates and children to 24 months |
| | | 49281-0574-88,<br>49281-0575-00<br>49281-0574-88 | Respiratory syncytial virus (RSV) monoclonal antibody, IgG1κ, (nirsevimab-alip), 1 mL, neonates and children to 24 months |
| Clesrovimab (ENFLONSIA®) | | 00006-5073-99,<br>00006-5073-02,<br>00006-5073-01 | Respiratory syncytial virus (RSV), monoclonal antibody, IgG1κ, (clesrovimab-cfor), 0.7 mL, neonate and infant, preservative free |
| Palivizumab (Synagis®) | 90378 | 66658-230-01,<br>66658-0230-01 | SYNAGIS- palivizumab injection, solution, 50 mg / 0.5 mL Single-Dose Vial |
| | | 66658-231-01,<br>66658-0231-01 | SYNAGIS- palivizumab injection, solution, 100 mg / 1.0 mL Single-Dose Vial |

Abbreviations: CPT, current procedural terminology; NDC, national drug codes; RSV, respiratory syncytial virus

* As identified by ACOG and AAFP (https://www.acog.org/practice-management/coding/coding-library/new-cpt-codes-for-rsv-vaccine-administration; https://www.aafp.org/pubs/fpm/blogs/gettingpaid/entry/rsv-antibody-vaccines.html).

**Table S2. Definition of exposure and mediator variables in CAUSAL-RSV analysis.**

| Variable | Role | Value | Definition |
|---|---|---|---|
| RSVpreF vaccine | Exposure | 1 = Yes<br>0 = No | NDC or CPT code consistent with RSVpreF vaccination (see Appendix B) in outpatient or inpatient claims during 32-36 weeks of pregnancy |
| Date of RSVpreF vaccine | Exposure | ####-##-## | Service date of RSVpreF vaccination in claims record |
| Gestational age at RSVpreF vaccine | Exposure | 32-36 | Week of gestation at RSVpreF vaccination, calculated as (vaccination date – last menstrual period)/7 |
| mAb administration | Mediator | 1 = Yes<br>0 = No | NDC or CPT code consistent with administration of monoclonal antibody in infant (see Appendix B) in inpatient or outpatient claims |
| mAb type | Mediator | 1 = Nirsevimab<br>2 = Clesrovimab | Type of monoclonal antibody based on NDC or CPT code |
| Date of mAb administration | Mediator | ####-##-## | Service date of monoclonal antibody administration in claims record |
| Infant age at mAb administration | Mediator | 0-24 | Infant age in weeks at first monoclonal antibody administration, calculated as (mab date – infant dob)/7 |

Abbreviations: CPT, Current Procedural Terminology; mAb, monoclonal antibody; NDC, National Drug Code

**Table S3. Logical Observation Identifiers Names and Codes (LOINC®) codes* for the identification of laboratory test results for respiratory syncytial virus.**

| Variable | Description |
|---|---|
| **Rapid tests** | |
| 68966-1 | Respiratory syncytial virus Ag [Presence] in Respiratory specimen by Immunoassay |
| 72885-7 | Respiratory syncytial virus Ag [Presence] in Nasopharynx by Immunoassay |
| 5876-8 | Respiratory syncytial virus Ag [Presence] in Specimen by Immunoassay |
| **NAAT Tests** | |
| 30075-6 | Respiratory syncytial virus A RNA [Presence] in Specimen by NAA with probe detection |
| 30076-4 | Respiratory syncytial virus B RNA [Presence] in Specimen by NAA with probe detection |
| 76089-2 | Respiratory syncytial virus RNA [Presence] in Specimen by NAA |
| 40988-8 | RSV RNA [Presence] in Specimen by Probe.amp.tar |
| 92131-2 | RSV RNA [Presence] in Respiratory specimen by Nucleic Acid Amplification (NAA). |
| 85479-4 | Respiratory syncytial virus RNA [Presence] in Upper respiratory specimen by NAA with probe detection |
| 82176-9 | Respiratory syncytial virus RNA [Presence] in Nasopharynx by NAA with non-probe detection |
| 77022-2 | Respiratory syncytial virus A RNA [Presence] in Nasopharynx by NAA with probe detection |
| 77023-0 | Respiratory syncytial virus B RNA [Presence] in Nasopharynx by NAA with probe detection |

Abbreviations: Ag, antigen; NAAT, nucleic acid amplification testing

*As identified by Centers for Disease Control and Prevention LOINC mapping (https://view.officeapps.live.com/op/view.aspx?src=https%3A%2F%2Fwww.cdc.gov%2Fcsels%2Fdls%2Fdocuments%2Flivd_test_code_mapping%2FLIVD-SARS-CoV-2-2024-11-14.xlsx&wdOrigin=BROWSELINK) and Find a Code (https://www.findacode.com/index.html).

**Table S4. Definition of primary and exploratory outcome variables in CAUSAL-RSV analysis.**

| Variable | Role | Value | Definition |
|---|---|---|---|
| Medically-attended RSV-associated LRTD hospitalization | Primary outcome | 1 = Yes<br>0 = No | Inpatient claim with an LRTD diagnosis (see Appendix C) AND a laboratory record with positive detection of RSV by rapid antigen or polymerase chain reaction (PCR) within 14 days before through 3 days after the LRTD diagnosis (see Table S5) |
| Date of first medically-attended RSV-associated LRTD | Primary outcome | Date | Date of first inpatient diagnosis from inpatient claims |
| Infant age (in weeks) at medically-attended RSV-associated LRTD hospitalization | Primary outcome | 0-32 | Age of infant in weeks at inpatient diagnosis from inpatient claims calculated as (diagnosis date – date of birth)/7 |
| Severe medically-attended RSV-associated LRTD hospitalization | Exploratory outcome | 1 = Yes<br>0 = No | Inpatient claim with an LRTD diagnosis (see Appendix C) AND requirement for mechanical ventilation and/or critical care AND a laboratory record with positive detection of RSV by rapid antigen or PCR within 14 days before through 3 days after the LRTD diagnosis (see Table S5) |
| Date of first severe medically-attended RSV-associated LRTD | Exploratory outcome | Date | Date of first inpatient diagnosis from inpatient claims |
| Infant age (in weeks) at severe medically-attended RSV-associated LRTD hospitalization | Exploratory outcome | 0-32 | Age of infant in weeks at inpatient diagnosis calculated as (diagnosis date – date of birth)/7 |
| Medically-attended RSV-associated LRTD | Exploratory outcome | 1 = Yes<br>0 = No | Inpatient or outpatient claim with an LRTD diagnosis (see Appendix C) AND a laboratory record with positive detection of RSV by rapid antigen or PCR within 14 days before through 3 days after the LRTD diagnosis (see Table S5) |
| Date of first medically-attended RSV-associated LRTD | Exploratory outcome | Date | Date of first inpatient or outpatient diagnosis from claim record |
| Infant age (in weeks) at medically-attended RSV-associated LRTD | Exploratory outcome | 0-32 | Age of infant in weeks at inpatient or outpatient diagnosis from claims calculated as (diagnosis date – date of birth)/7 |
| All-cause LRTD hospitalization | Exploratory outcome | 1 = Yes<br>0 = No | Inpatient claim with an LRTD diagnosis (see Table S5) |
| Date of all-cause LRTD hospitalization | Exploratory outcome | Date | Date of first inpatient LRTD diagnosis from claims record |
| Infant age (in weeks) at all-cause LRTD hospitalization | Exploratory outcome | 0-32 | Age of infant in weeks at inpatient LRTD diagnosis from claims calculated as (diagnosis date – date of birth)/7 |

Abbreviations: LRTD, lower respiratory tract disease; PCR, polymerase chain reaction

**Table S5. International Classification of Diseases (10th edition, clinical modification; ICD-10-CM) codes* to identify a diagnosis of lower respiratory tract disease.**

| Code | Description |
|---|---|
| B974 | Respiratory syncytial virus as the cause of diseases classified elsewhere |
| J1100 | Influenza with pneumonia, virus not identified |
| J1108 | Influenza with other manifestations, virus not identified |
| J111 | Influenza with other respiratory manifestations, virus not identified |
| J118 | Influenza with other manifestations, virus not identified |
| J121 | Respiratory syncytial virus pneumonia |
| J1289 | Viral pneumonia, unspecified |
| J129 | Viral pneumonia, unspecified |
| J168 | Pneumonia due to other specified infectious organisms |
| J17 | Pneumonia diseases classified elsewhere |
| J180 | Bronchopneumonia, unspecified organism |
| J181 | Lobar pneumonia, unspecified organism |
| J182 | Hypostatic pneumonia, unspecified organism |
| J188 | Other pneumonia, organism unspecified |
| J189 | Pneumonia, unspecified organism |
| J205 | Acute bronchitis due to respiratory syncytial virus |
| J208 | Acute bronchitis due to other specified organisms |
| J209 | Acute bronchitis, unspecified |
| J210 | Acute bronchiolitis due to respiratory syncytial virus |
| J218 | Acute bronchiolitis due to other specified organisms |
| J219 | Acute bronchiolitis, unspecified |
| J40 | Bronchitis, not specified as acute or chronic |
| J22 | Unspecified acute lower respiratory infection |
| J410 | Simple chronic bronchitis |
| J411 | Mucopurulent chronic bronchitis |
| J418 | Mixed simple and mucopurulent chronic bronchitis |
| J42 | Unspecified chronic bronchitis |
| J440 | Chronic obstructive pulmonary disease with (acute) lower respiratory infection |
| J441 | Chronic obstructive pulmonary disease with (acute) exacerbation |
| J449 | Chronic obstructive pulmonary disease, unspecified |
| J4520 | Mild intermittent asthma, uncomplicated |
| J4521 | Mild intermittent asthma with (acute) exacerbation |
| J4522 | Mild intermittent asthma with status asthmaticus |
| J4530 | Mild persistent asthma, uncomplicated |
| J4531 | Mild persistent asthma with (acute) exacerbation |
| J4532 | Mild persistent asthma with status asthmaticus |
| J4540 | Moderate persistent asthma, uncomplicated |
| J4541 | Moderate persistent asthma with (acute) exacerbation |
| J4542 | Moderate persistent asthma with status asthmaticus |
| J4550 | Severe persistent asthma, uncomplicated |
| J4551 | Severe persistent asthma with (acute) exacerbation |
| J4552 | Severe persistent asthma with status asthmaticus |
| J4590 | Unspecified asthma |
| J45901 | Unspecified asthma with (acute) exacerbation |
| J45902 | Unspecified asthma with status asthmaticus |
| J45909 | Unspecified asthma, uncomplicated |
| J4599 | Other asthma |
| J45990 | Exercise induced bronchospasm |
| J45991 | Cough variant asthma |
| J45998 | Other asthma |

* As identified by Gatenberg et al. Risk analysis of respiratory syncytial virus among infants in the United States by birth month. J Pediatr Infect Dis Soc 2024; 13(6): 317-27.

**Table S6. Definition of covariates in CAUSAL-RSV analysis.**

| Variable | Role | Value | Definition |
|---|---|---|---|
| **Maternal / Household Covariates** | | | |
| Maternal age | Demographic covariate | 18-49 | Mother's age at LMP based on enrollment data |
| Household income | Demographic covariate | 1 = <$40,000<br>2 = $40,000 to $74,999<br>3 = $75,000 to $124,999<br>4 = $125,000 to $199,999<br>5 = $200,000+ | Household income based on enrollment data |
| Educational attainment | Exposure | 1 = Less than 12th grade<br>2 = HS diploma<br>3 = Less than bachelor's degree<br>4 = Bachelor's degree or higher | Area-level educational attainment based on enrollment data |
| Smoking | Health covariate | 1 = Yes<br>0 = No | Any record of ICD-10CM diagnosis of smoking during 365 days prior to LMP through date of pregnancy end (see Table S7) |
| Asthma | Health covariate | 1 = Yes<br>0 = No | ICD-10-CM diagnosis of asthma during 365 days prior to LMP (see Table S7) |
| Other chronic lung disease | Health covariate | 1 = Yes<br>0 = No | ICD-10-CM diagnosis of other chronic lung disease during 365 days prior to LMP (see Table S7) |
| Immunocompromise (including solid organ transplant) | Health covariate | 1 = Yes<br>0 = No | ICD-10-CM diagnosis of immunocompromise or immunosuppressing conditions during 365 days prior to LMP (see Table S7) |
| Pre-existing diabetes | Health covariate | 1 = Yes<br>0 = No | ICD-10-CM diagnosis of pre-existing diabetes during 365 days prior to LMP |
| Essential hypertension | Health covariate | 1 = Yes<br>0 = No | ICD-10-CM diagnosis of pre-existing hypertension during 365 days prior to LMP (see Table S7) |
| Chronic kidney disease | Health covariate | 1 = Yes<br>0 = No | ICD-10-CM diagnosis of chronic kidney disease during 365 days prior to LMP (see Table S7) |
| Chronic heart disease | Health covariate | 1 = Yes<br>0 = No | ICD-10-CM diagnosis of chronic heart disease during 365 days prior to LMP (see Table S7) |
| Obesity | Health covariate | 1 = Yes<br>0 = No | ICD-10-CM diagnosis of obesity during 365 days prior to LMP (see Table S7) |
| Gestational diabetes | Health covariate | 1 = Yes<br>0 = No | ICD-10-CM diagnosis of gestational diabetes between LMP and vaccination date (see Table S7) |
| Gestational hypertension | Health covariate | 1 = Yes<br>0 = No | ICD-10-CM diagnosis of gestational hypertension between LMP and vaccination date (see Table S7) |

| | | | |
|---|---|---|---|
| Pre-eclampsia | Health covariate | 1 = Yes<br>0 = No | ICD-10-CM diagnosis of pre-eclampsia between LMP and vaccination date (see Table S7) |
| HELLP / Eclampsia | Health covariate | 1 = Yes<br>0 = No | ICD-10-CM diagnosis of HELLP/eclampsia between LMP and vaccination date (see Table S7) |
| Received Tdap vaccine | Health covariate | 1 = Yes<br>0 = No | CPT or NDC code indicating receipt of Tdap vaccine between LMP and vaccination date (see Table S7) |
| Received influenza vaccine | Health covariate | 1 = Yes<br>0 = No | CPT or NDC code indicating receipt of influenza vaccine between LMP and vaccination date (see Table S7) |
| Received COVID-19 vaccine | Health covariate | 1 = Yes<br>0 = No | CPT or NDC code indicating receipt of a COVID-19 vaccine between LMP and vaccination date (see Table S7) |
| **Infant Covariates** | | | |
| Infant sex | Demographic covariate | 1 = Male<br>2 = Female | Infant sex based on infant enrollment data |
| Prematurity | Health covariate | 1 = Yes<br>0 = No | Infant born at gestational age 20 to 36 weeks based on estimated gestational age from pregnancy algorithm or ICD-10-CM diagnoses (see Table S8) |
| Low birthweight | Health covariate | 1 = Yes<br>0 = No | Infant born low birthweight based on ICD-10-CM diagnoses (see Table S8) |
| Small-for-gestational age | Health covariate | 1 = Yes<br>0 = No | Infant born small-for-gestational age based on ICD-10-CM diagnoses (see Table S8) |
| Trisomy 21 | Health covariate | 1 = Yes<br>0 = No | Infant born with a diagnosed congenital anomaly based on ICD-10-CM diagnoses (see Table S8) |
| Cesarean delivery | Health covariate | 1 = Yes<br>0 = No | Infant born by cesarean delivery based on ICD-10-CM and CPT codes (see Table S8) |
| Neonatal intensive care unit (NICU) admission | Health covariate | 1 = Yes<br>0 = No | Infant admitted to NICU based on the presence of any CPT code during 0-28 days after birth (see Table S8) |
| Chronic lung disease of prematurity | Health covariate | 1 = Yes<br>0 = No | Infant diagnosed with chronic lung disease based on ICD-10-CM diagnoses (see Table S8) |
| Congenital heart disease | Health covariate | 1 = Yes<br>0 = No | Infant diagnosed with congenital heart disease based on ICD-10-CM diagnoses (see Table S8) |
| Neuromuscular disease impairing respiratory function | Health covariate | 1 = Yes<br>0 = No | Infant diagnosed with neuromuscular disease based on ICD-10-CM diagnoses from birth to end of follow-up (see Table S8) |
| Immunocompromising conditions | Health covariate | 1 = Yes<br>0 = No | Infant diagnosed with immunocompromising condition based on ICD-10-CM diagnoses from birth to end of follow-up (see Table S8) |

Abbreviations: CPT, Current Procedural Terminology; HIV, human immunodeficiency virus; ICD-10-CM, International Classification of Diseases (10th edition, clinical modification; ICD-10-CM); NDC, National Drug Codes.

**Table S7. International Classification of Diseases (10th edition, clinical modification; ICD-10-CM), Current Procedural Terminology, and National Drug Codes to identify baseline maternal covariates.**

| Variable | Description |
|---|---|
| **Health covariates** | **ICD-10-CM code** |
| Smoking | F17210, F17213, F17218, F17219, F17220, F17223, F17228, F17229, F17290, F17293, F17298, F17299, T65211A, T65211D, T65211S, T65212A, T65212D, T65212S, T65213A, T65213D, T65213S, T65214A, T65214D, T65214S, Z716, Z720, Z87891 |
| Asthma | J45902, J45901, J4552, J4551, J4550, J455, J4542, J4541, J4540, J454, J4532, J4531, J4530, J453, J4522, J4521, J452, J45 |
| Other chronic lung disease | E84, E840, E841, E8411, E8419, E848, E849, J8410, J84112, I2723 |
| Immunocompromised (including solid organ transplant) | B20, O987, O9871, O98711, O98712, O98713, O98719, O9872, O9873, D80, D800, D801, D802, D803, D804, D805, D806, D807, D808, D809, D81, D810, D811, D812, D8130, D8131, D8132, D8139, D814, D815, D816, D817, D81810, D81818, D81819, D8182, D8189, D819, D82, D820, D821, D822, D823, D824, D828, D829, D83, D830, D831, D832, D838, D839, D84, D840, D841, D8481, D84821, D84822, D8489, D849, D89, D890, D891, D892, D893, D8940, D8941, D8942, D8943, D8944, D8949, D89810, D89811, D89812, D89813, D8982, D89831, D89832, D89833, D89834, D89835, D89839, D8989, D899, D47OZ1, T451X1, T451X1A, T451X1s, Z21, Z510, Z511, Z5111, Z5112, Z795, Z7951, Z7952, Z48290, Z9481, Z9484, Z482, Z4821, Z4822, Z4823, Z4824, Z4828, Z48280, Z48288, Z4829, Z48298, Z940, Z941, Z942, Z943, Z944, Z945, Z946, Z947, Z948, Z9482, Z9483, Z9489, Z949 |
| Pre-existing diabetes | E08, E080, E0800, E0801, E081, E0810, E0811, E082, E0821, E0822, E0829, E083, E0831, E08311, E08319, E0832, E08321, E083211, E083212, E083213, E083219, E08329, E083291, E083292, E083293, E083299, E0833, E08331, E083311, E083312, E083313, E083319, E08339, E083391, E083392, E083393, E083399, E0834, E08341, E083411, E083412, E083413, E083419, E08349, E083491, E083492, E083493, E083499, E0835, E08351, E083511, E083512, E083513, E083519, E08352, E083521, E083522, E083523, E083529, E08353, E083531, E083532, E083533, E083539, E08354, E083541, E083542, E083543, E083549, E08355, E083551, E083552, E083553, E083559, E08359, E083591, E083592, E083593, E083599, E0836, E0837, E0837X1, E0837X2, E0837X3, E0837X9, E0839, E084, E0840, E0841, E0842, E0843, E0844, E0849, E085, E0851, E0852, E0859, E086, E0861, E08610, E08618, E0862, E08620, E08621, E08622, E08628, E0863, E08630, E08638, E0864, E08641, E08649, E0865, E0869, E088, E089, E09, E090, E0900, E0901, E091, E0910, E0911, E092, E0921, E0922, E0929, E093, E0931, E09311, E09319, E0932, E09321, E093211, E093212, E093213, E093219, E09329, E093291, E093292, E093293, E093299, E0933, E09331, E093311, E093312, E093313, E093319, E09339, E093391, E093392, E093393, E093399, E0934, E09341, E093411, E093412, E093413, E093419, E09349, E093491, E093492, E093493, E093499, E0935, E09351, E093511, E093512, E093513, E093519, E09352, E093521, E093522, E093523, E093529, E09353, E093531, E093532, E093533, E093539, E09354, E093541, E093542, E093543, E093549, E09355, E093551, E093552, E093553, E093559, E09359, E093591, E093592, E093593, E093599, E0936, E0937, E0937X1, E0937X2, E0937X3, E0937X9, E0939, E094, E0940, E0941, E0942, E0943, E0944, E0949, E095, E0951, E0952, E0959, E096, E0961, E09610, E09618, E0962, E09620, E09621, E09622, E09628, E0963, E09630, E09638, E0964, E09641, E09649, E0965, E0969, E098, E099, E10, E101, E1010, E1011, E102, E1021, E1022, E1029, E103, E1031, E10311, E10319, E1032, E103211, E103212, E103213, E103219, E10329, E103291, E103292, E103293, E103299, E1033, E10331, E103311, E103312, E103313, E103319, E10339, E103391, E103392, E103393, E103399, E1034, E10341, E103411, E103412, E103413, E103419, E10349, E103491, E103492, E103493, E103499, E1035, E10351, E103511, E103512, E103513, E103519, E10352, E103521, E103522, E103523, E103529, E10353, E103531, E103532, E103533, E103539, E10354, E103541, E103542, E103543, E103549, E10355, E103551, E103552, E103553, E103559, E10359, E103591, E103592, E103593, E103599, E1036, E1037, E1037X1, E1037X2, E1037X3, E1037X9, E1039, E104, E1040, E1041, E1042, E1043, E1044, E1049, E105, E1051, E1052, E1059, E106, E1061, E10610, E10618, E1062, E10620, E10621, E10622, E10628, E1063, E10630, E10638, E1064, E10641, E10649, E1065, E1069, E108, E109, E11, E110, E1100, E1101, E111, E1110, E1111, E112, E1121, |

| | |
|---|---|
| | E1122, E1129, E113, E1131, E11311, E11319, E1132, E113211, E113212, E113213, E113219, E11329, E113291, E113292, E113293, E113299, E1133, E11331, E113311, E113312, E113313, E113319, E11339, E113391, E113392, E113393, E113399, E1134, E11341, E113411, E113412, E113413, E113419, E11349, E113491, E113492, E113493, E113499, E1135, E11351, E113511, E113512, E113513, E113519, E11352, E113521, E113522, E113523, E113529, E11353, E113531, E113532, E113533, E113539, E11354, E113541, E113542, E113543, E113549, E11355, E113551, E113552, E113553, E113559, E11359, E113591, E113592, E113593, E113599, E1136, E1137, E1137X1, E1137X2, E1137X3, E1137X9, E1139, E114, E1140, E1141, E1142, E1143, E1144, E1149, E115, E1151, E1152, E1159, E116, E1161, E11610, E11618, E1162, E11620, E11621, E11622, E11628, E1163, E11630, E11638, E1164, E11641, E11649, E1165, E1169, E118, E119, E13, E130, E1300, E1301, E131, E1310, E1311, E132, E1321, E1322, E1329, E133, E1331, E13311, E13319, E1332, E13321, E133211, E133212, E133213, E133219, E13329, E133291, E133292, E133293, E133299, E1333, E13331, E133311, E133312, E133313, E133319, E13339, E133391, E133392, E133393, E133399, E1334, E13341, E133411, E133412, E133413, E133419, E13349, E133491, E133492, E133493, E133499, E1335, E13351, E133511, E133512, E133513, E133519, E13352, E133521, E133522, E133523, E133529, E13353, E133531, E133532, E133533, E133539, E13354, E133541, E133542, E133543, E133549, E13355, E133551, E133552, E133553, E133559, E13359, E133591, E133592, E133593, E133599, E1336, E1337, E1337X1, E1337X2, E1337X3, E1337x9, E1339, E134, E1340, E1341, E1342, E1343, E1344, E1349, E135, E1351, E1352 , E1359, E136, E1361, E13610, E13618, E1362, E13620, E13621, E13622, E13628, E1363, E13630, E13638, E1364, E13641, E13649, E1365, E1369, E138, E139, E15 |
| Essential hypertension | I10, I15, I150, I151, I152, I158, I159, I16, I160, I161, I169, I674, O10, O100, O1001, O10011, O10012, O10013, O10019, O1002, O1003, O109, O1091, O10911, O10912, O10913, O10919, O1092, O1093 |
| Chronic kidney disease | N18, N183, N1830, N1831, N1832, N184, N185, N186, N189, N19, Z49, Z490, Z4901, Z4902, Z493, Z4931, Z4932, Z9115, Z940_kidney, Z992 |
| Chronic heart disease | I42, I420, I421, I422, I423, I424, I425, I426, I427, I428, I429, I43, I50, I501, I502, I5020, I5021, I5022, I5023, I503, I5030, I5031, I5032, I5033, I504, I5040, I5041, I5042, I5043, I508, I5081, I50810, I50811, I50812, I50813, I50814, I5082, I5083, I5084, I5089, I509 |
| Obesity | E6601, E6609, E661, E662, E668, E669, E6830, E6831, E6832, E6833, E6834, E6835, E6836, E6837, E6838, E6839, E6841, E6842, E6843, E6844, E6845, E6854 |
| Gestational diabetes | O24410, O24414, O24415, O24419, O24811, O24812, O24813, O24819, O24911, O24912, O24913, O24919 |
| Gestational hypertension* | O131, O132, O133, O139 |
| Pre-eclampsia* | O111, O112, O113, O119, O1400, O1402, O1403, O1410, O1412, O1413, O1490, O1492, O1493 |
| HELLP / Eclampsia* | O1420, O1422, O1423, O1500, O1502, O1503 |

| **Immunization covariates** | **CPT codes** | **NDC codes** |
|---|---|---|
| Tdap vaccine | 90715 | 49281-0400-05, 49281-0400-10, 49281-0400-20, 49281-0400-58, 49281-0400-89, 58160-0842-01, 58160-0842-00 |
| Influenza vaccine | 90653, 90656, 90657, 90658, 90660, 90661, 90662, 90672, 90673, 90674, 90682, 90686, 90687, 90688, 90756 | 58160-0909-52, 58160-0912-52, 19515-0814-52, 19515-0810-52, 49281-0423-50, 49281-0424-50, 33332-0323-03, 33332-0024-03, 33332-0423-10, 33332-0124-11, 49281-0639-15, 49281-0641-15, 49281-0123-65, 49281-0124-65, 49281-0125-65, 70461-0123-03, 70461-0024-03, 70461-0025-03, 70461-0323-03, 70461-0423-10, 70461-0654-03, 70461-0554-10, 49281-0723-10, 49281-0724-10, 49281-0725-10, 66019-0310-10, 66019-0311-10, 58160-0909-41, 58160-0912-41, 19515-0814-41, 19515-0810-41, 49281-0423-88, |

| | | |
|---|---|---|
| | | 49281-0424-88, 49281-0123-88, 49281-0124-88, 49281-0125-88, 49281-0723-88, 49281-0724-88, 49281-0725-88, 70461-0123-04, 70461-0323-04, 70461-0654-04 |
| COVID-19 vaccine | 91318, 91319, 91320, 91321, 91322, 91304, 90480 | 00069-2362-10, 00069-2362-01, 00069-2392-10, 00069-2392-01, 80777-0102-95, 80777-0102-04, 80777-0102-01, 80631-0105-02, 80631-0105-01, 00069-2432-01, 59267-4438-02, 59267-4438-01, 59267-4426-02, 59267-4426-01, 80777-0110-93, 80777-0110-01, 80777-0291-80, 80777-0291-09, 80777-0291-81, 59267-4331-02, 59267-4331-01, 59267-4315-02, 59267-4315-01, 80777-0102-96, 80777-0102-93, 80777-0287-92, 80777-0287-07, 0069-2432-10, 80777-0110-96, 80631-0107-01, 80631-0107-10, 0069-2528-10, 00069-2528-01, 00069-2501-10, 00069-2501-01, 80777-0112-96, 80777-0112-01, 80777-0112-93, 80777-0112-88, 80777-0400-60, 80777-0400-17<br>80777-0400-61, 80777-0400-62, 80777-0113-80, 80777-0113-09<br>80777-0113-87, 0631-0207-10, 80631-0207-01 |

Abbreviations: CPT, Current Procedural Terminology; ICD-10-CM, International Classification of Diseases (10th edition, clinical modification; ICD-10-CM); NDC, National Drug Codes.

*Codes are restricted to those that would be identified at or before 32 weeks of pregnancy.

**Table S8. International Classification of Diseases (10th edition, clinical modification; ICD-10-CM) and Current Procedural Terminology (CPT) codes to measure infant health covariates.**

| Variable | ICD-10-CM / CPT codes |
|---|---|
| **Prematurity** | **ICD-10-CM: P0720, P0721, P0722, P0723, P0724, P0725, P0726, P0729, P0730, P0731, P0732, P0733, P0734, P0735, P0736, P0737, P0738, P0739** |
| **Low birthweight** | **ICD-10-CM: P0700, P0701, P0702, P0703, P0704, P0705, P0706, P0707, P0708, P0709, P0710, P0711, P0712, P0713, P0714, P0715, P0716, P0717, P0718, P0719** |
| **Small-for-gestational age** | **ICD-10-CM: P0510, P0511, P0512, P0513, P0514, P0515, P0516, P0517, P0518, P0519, P059** |
| **Trisomy 21** | **ICD-10-CM: Q900, Q901, Q902, Q909** |
| **Cesarean delivery** | **ICD-10-CM: O82, Z3701, Z3721, Z3731, Z3741, Z379**<br><br>**CPT: 59510, 59514, 59515, 59525** |
| **NICU admission** | **CPT: 99468, 99469, 99477, 99478, 99479** |
| **Chronic lung disease of prematurity** | **ICD-10-CM: P270, P271, P278, P279** |
| Wilson-Mikity syndrome | ICD-10-CM: P270 |
| Bronchopulmonary dysplasia | ICD-10-CM: P271 |
| Other or unspecified chronic respiratory disease originating during perinatal period | ICD-10-CM: P278, P279 |
| **Congenital heart disease** | **ICD-10-CM: Q200, Q201, Q202, Q203, Q204, Q205, Q206, Q208, Q209, Q210, Q2110, Q2111, Q2112, Q2113, Q2114, Q2115, Q2116, Q2119, Q2120, Q2121, Q2122, Q2123, Q213, Q214, Q218, Q219, Q220, Q221, Q222, Q223, Q224, Q225, Q226, Q228, Q229, Q230, Q231, Q232, Q233, Q234, Q2381, Q2382, Q2388, Q239, Q240, Q241, Q242, Q243, Q244, Q245, Q246, Q248, Q249, Q250, Q251, Q2521, Q2529, Q253, Q2540, Q2541, Q2542, Q2543, Q2544, Q2545, Q2546, Q2547, Q2548, Q2549, Q255, Q256, Q2571, Q2572, Q2579, Q258, Q259, Q260, Q261, Q262, Q263, Q264, Q265, Q266, Q268, Q269** |
| Congenital malformations of cardiac chambers and connections | ICD-10-CM: Q200, Q201, Q202, Q203, Q204, Q205, Q206, Q208, Q209 |
| Congenital malformations of cardiac septa | ICD-10-CM: Q210, Q2110, Q2111, Q2112, Q2113, Q2114, Q2115, Q2116, Q2119, Q2120, Q2121, Q2122, Q2123, Q213, Q214, Q218, Q219 |
| Congenital malformations of pulmonary and tricuspid valves | ICD-10-CM: Q220, Q221, Q222, Q223, Q224, Q225, Q226, Q228, Q229 |
| Congenital malformations of aortic and mitral valves | ICD-10-CM: Q230, Q231, Q232, Q233, Q234, Q2381, Q2382, Q2388, Q239 |
| Other congenital malformations of heart | ICD-10-CM: Q240, Q241, Q242, Q243, Q244, Q245, Q246, Q248, Q249 |
| Congenital malformations of great arteries | ICD-10-CM: Q250, Q251, Q2521, Q2529, Q253, Q2540, Q2541, Q2542, Q2543, Q2544, Q2545, Q2546, Q2547, Q2548, Q2549, Q255, Q256, Q2571, Q2572, Q2579, Q258, Q259 |
| Congenital malformations of great veins | ICD-10-CM: Q260, Q261, Q262, Q263, Q264, Q265, Q266, Q268, Q269 |
| **Neuromuscular disease** | **ICD-10-CM: G120, G121, G1220, G1221, G1222, G1223, G1224, G1225, G1229, G128, G129, G7000, G7001, G7002, G702, G7100, G7101, G7102, G7103, G71031, G71032, G71033, G710340, G710341, G710342, G710349, G71035, G71036, G71038, G71039, G7109, G7111, G7112, G7113, G7119, G7120, G7121, G71220, G71228, G7129, G713, G718, G719, P940, P942** |
| Spinal muscular atrophies & motor neuron diseases | ICD-10-CM: G120, G121, G1220, G1221, G1222, G1223, G1224, G1225, G1229, G128, G129 |
| Myasthenia gravis & neuromuscular junction disorders | ICD-10-CM: G7000, G7001, G7002 |

| | |
|---|---|
| Congenital myasthenia | ICD-10-CM: G702 |
| Muscular dystrophy & primary muscular disorders | ICD-10-CM: G7100, G7101, G7102, G7103, G71031, G71032, G71033, G710340, G710341, G710342, G710349, G71035, G71036, G71038, G71039, G7109, G7111, G7112, G7113, G7119, G7120, G7121, G71220, G71228, G7129, G713, G718, G719 |
| Congenital hypotonia | ICD-10-CM: P940, P942 |
| **Immunocompromising conditions** | **ICD-10-CM: D800, D801, D802, D803, D804, D805, D806, D807, D808, D809, D8130, D8131, D8132, D8139, D814, D815, D816, D817, D81810, D81818, D81819, D8182, D8189, D819, D820, D8221, D822, D823, D824, D828, D829, D830, D831, D832, D838, D839, D840, D841, D8481, D84821, D84822, D8489, D849, B200, B201, B202, B203, B204, B205, B206, B207, B208, B209, B210, B211, B212, B213, B217, B218, B219, B220, B221, B222, B227, B230, B231, B231, B232, Z206** |
| Primary immunodeficiency | ICD-10-CM: D800, D801, D802, D803, D804, D805, D806, D807, D808, D809, D8130, D8131, D8132, D8139, D814, D815, D816, D817, D81810, D81818, D81819, D8182, D8189, D819, D820, D8221, D822, D823, D824, D828, D829, D830, D831, D832, D838, D839, D840, D841, D8481, D84821, D84822, D8489, D849 |
| HIV (or maternal exposure to HIV) | ICD-10-CM: B200, B201, B202, B203, B204, B205, B206, B207, B208, B209, B210, B211, B212, B213, B217, B218, B219, B220, B221, B222, B227, B230, B231, B231, B232, Z206 |

Abbreviations: CPT, Current Procedural Terminology; HIV, human immunodeficiency virus; ICD-10-CM, International Classification of Diseases (10th edition, clinical modification; ICD-10-CM); NICU, neonatal intensive care unit.

**Figure S1. Respiratory syncytial virus (RSV) activity used to establish maternal-infant eligibility criteria.**

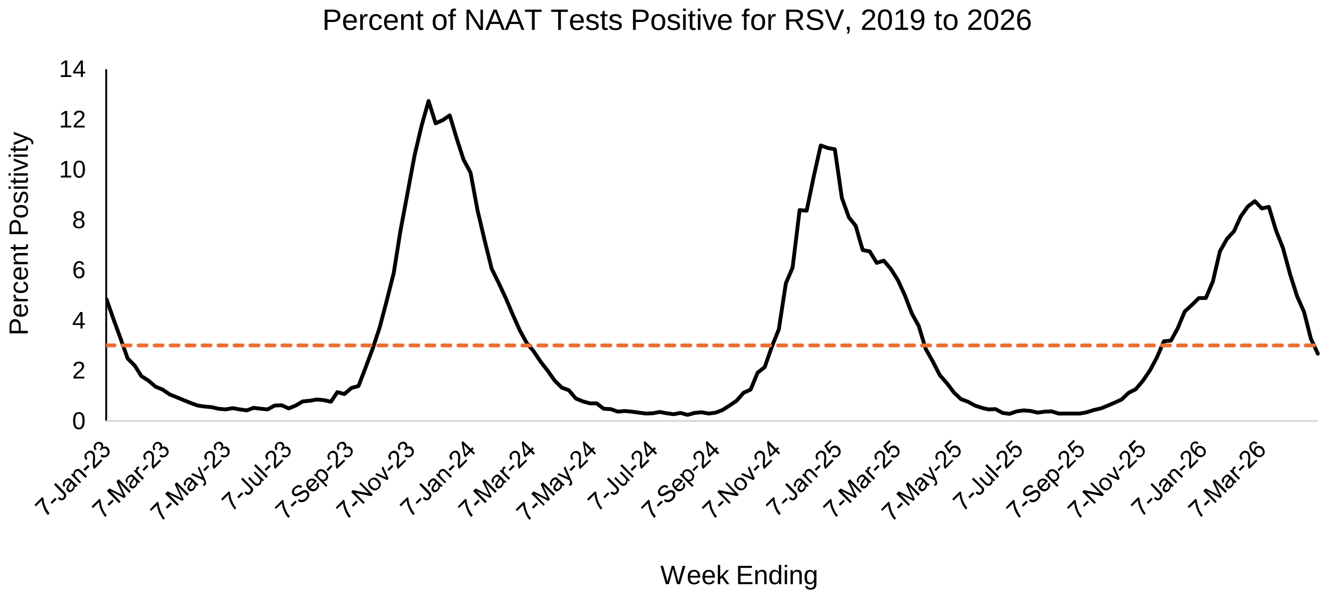


| RSV Season | RSV Season Dates† | Infant DOB§ | LMP criteria¶ |
|---|---|---|---|
| 2023-24 Season | October 7, 2023 to March 23, 2024 | February 7, 2023 to March 22, 2024 | January 20, 2023 to June 21, 2023 |
| 2024-25 Season | November 9, 2024 to April 19, 2025 | March 9, 2024 to April 19, 2025 | January 20, 2024 to June 21, 2024 |
| 2025-26 Season | November 29, 2025 to April 25, 2026 | March 29, 2025 to April 25, 2026 | January 20, 2025 to June 21, 2025 |

* National estimates of the percent of NAAT tests positive for RSV by surveillance week were obtained from the National Respiratory and Enteric Virus Surveillance System (Source: CDC, https://www.cdc.gov/nrevss/php/dashboard/index.html).

† Season start and end dates were defined based on the weeks during which the percent of NAAT tests that were positive for RSV was ≥3% (as per: https://www.cdc.gov/nrevss/php/dashboard/index.html; https://www.cdc.gov/mmwr/volumes/72/wr/mm7214a1.htm).

§ To ensure eligible infants were at risk of the study outcome, we included only those either born during the RSV season or <8 months old at the start of the RSV season.

¶LMP criteria were established based on being no less than 32 weeks gestational age at the start of RSV immunization (September 1) and no more than 32 weeks gestational age at the end of typical RSV immunization (January 31). Vaccine availability was estimated based on distribution data in VSD (source: https://www.cdc.gov/rsvvaxview/dashboard/pregnant-women-coverage.html).
